# High *Mycobacterium bovis* exposure but low IGRA positivity in UK farm workers

**DOI:** 10.1101/2024.06.27.24309580

**Authors:** Amy Thomas, Alice Halliday, Genevieve Clapp, Ross Symonds, Noreen Hopewell-Kelley, Carmel McGrath, Lucy Wheeler, Anna Dacey, Nigel Noel, Andrea Turner, Isabel Oliver, James Wood, Ed Moran, Paul Virgo, Anu Goenka, Ellen Brooks-Pollock

## Abstract

**Background:** Between 1999 and 2021, 505 culture-confirmed cases of *M. bovis* disease in humans (zoonotic tuberculosis, TB) were identified in England. We aimed to estimate the prevalence of *M. bovis* infection in persons exposed to TB-infected cattle in England and identify any risk factors associated with latent TB infection (LTBI) in this population.

**Methods:** We co-developed a retrospective cohort study in southwest England, a bovine TB high risk area, with members of the UK farming community. A questionnaire captured participant characteristics, behaviours and farming practices. Linkage with historical herd testing data was used to categorise participants as low, medium or high risk for TB exposure. Interferon gamma release assay (IGRA) positivity with Quantiferon was used to determine LTBI status and linked to questionnaire data.

**Results:** We recruited 90 participants at agricultural shows and a standalone event. Participants were farmers/farm workers (79/90) and veterinary professionals (11/90). Median age was 45.5 years (IQR: 19–77); 58% were male; 66% reported BCG vaccination. *M. bovis* exposure was via direct contact with infected cattle and consumption of raw milk. One participant in the high-risk group was IGRA positive, all other participants were IGRA negative. Estimated IGRA positivity rate was 1.1% (95% CI 0.058%–7.0%) in all participants and 4.0% (95%CI 0.21%–22%) in participants with high exposure levels.

**Conclusions:** We found limited LTBI in individuals in contact with TB-infected cattle in England, despite high and prolonged exposure. We identified a high-risk group of farmers who should be prioritised for future engagement.

## Introduction

Zoonotic tuberculosis (TB) is a form of TB disease in humans caused primarily by infection with *Mycobacterium bovis (M. bovis)*, the main cause of TB in cattle. Transmission of *M. bovis* to humans from cattle can occur through consumption of contaminated unpasteurised milk and milk products, raw meat or through aerosols and human-to-human transmission can also occur. Cases of zTB predominate in low-income countries(1). Communities at risk of zTB tend to be rural, isolated populations, underserved by healthcare and cases are often not recorded(2). Once diagnosed, zoonotic TB is challenging to treat as *M. bovis* is resistant to pyrazinamide, a first line TB treatment.

In the UK, *M. bovis* disease in humans has largely been controlled through milk pasteurisation and culling infected cattle. Case reports of *M. bovis* disease in UK farmers have been documented(3). Between 1999 and 2021, 505 culture-confirmed cases of *M. bovis* disease in humans were identified in England, with numbers increasing until 2017(4)(fig 1). This represents a small proportion (∼1%) of all human TB cases, however *M. bovis* cases in England are demographically distinct from *M. tb* cases. *M. bovis* cases are more likely to be over 65 years of age and UK-born(5). Moreover, it is estimated that the strongest risk factor for human *M. bovis* infection in the UK is working in an agricultural or animal-related occupation (adjusted odds ratio 29.5, 95% CI: 16.9–51.6)(5).

**Figure 1:**
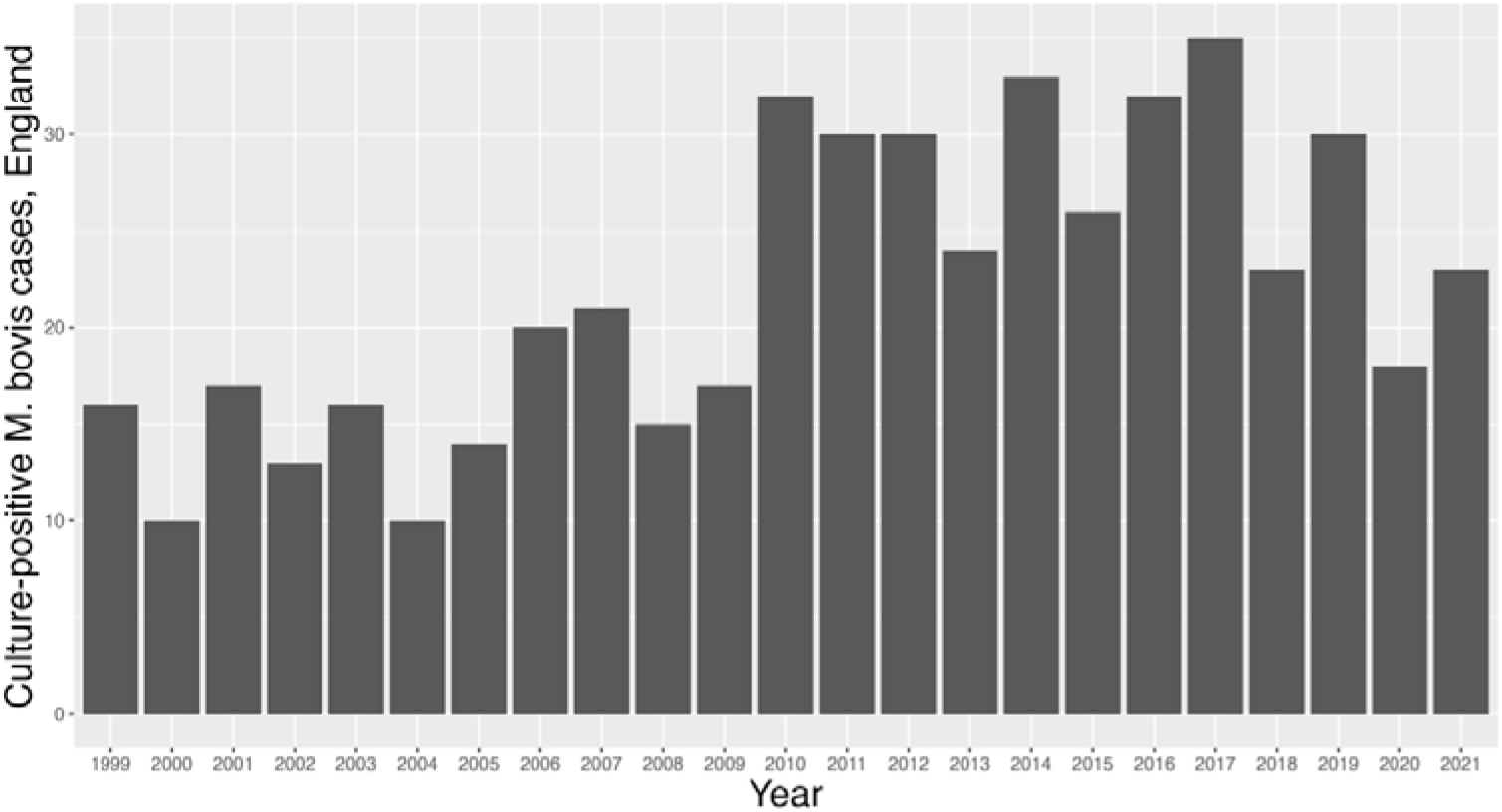
The number of culture-positive M. bovis cases in people diagnosed in England between 1999 and 2021. Data taken from UKHSA TB reports(4).

Bovine TB in cattle caused by *M. bovis* is a major problem for UK farming. In England, herds under movement restrictions due to TB increased from <1% at the end of 1996 to 11.3% at the end of 2011, including 1 in 5 herds in England’s high-risk areas(6). In 2017 and 2018, in excess of 30,000 cattle were slaughtered due to TB. Since then, incidence has decreased to ∼7% with 22,084 cattle slaughtered due to TB in 2022. Linking trends in incidence in cattle and humans is challenging because of the long natural history associated with TB disease. Individuals can develop disease years or decades following infection. In this study, we sought to estimate the prevalence of zTB infection in persons occupationally exposed to TB-infected cattle in southwest England. Furthermore, if feasible we aimed to identify what risk factors, if any, are associated with a positive IGRA in this population.

## Methods

### 1. Public Involvement

Public involvement and engagement (PPIE) involves working in partnership with people affected by research to ensure its relevance, acceptability, and accessibility(7). We involved members of the farming community throughout our study to co-develop feasibility, study design, materials and recruitment(8).

The first PPIE session, conducted prior to study funding, involved four farmers recruited through a veterinary practice in southwest England. Topics discussed included zTB, BCG vaccination, unpasteurised milk consumption and the acceptability of requesting a blood sample. Participants were remunerated for their time. A second PPIE session was conducted with a group of 22 farmers, with responses on the same topics captured anonymously using polling software. No remuneration was given at this session as it was part of a larger event co-organised with a local veterinary practice.

A third group PPIE session was conducted after receiving study funding. The session was online due to the COVID-19 pandemic and involved four farmers based in southwest England recruited via Twitter. The purpose of this session was to identify potential study locations that overcame the challenge of recruiting multiple farmers in a single location where a phlebotomist was based. This group identified agricultural shows as locations that agricultural workers and their families visit during their leisure time hence could have time to take part in the study. Participants were remunerated for their time.

We recruited one farmer from the third PPIE session to co-develop the study further. They were involved in study design, reviewing study documentation, recruitment via online farming forums and in-person at agricultural shows. The farmer co-developer was paid for his time on the project.

Our final PPIE session was aimed specifically at increasing the engagement of younger farmers with the study. This was conducted online with three young farmers, recruited via Facebook. This session identified the importance of farming social influencers, e.g. Jeremy Clarkson. Participants were remunerated for their time.

### 2. Study design

We conducted a retrospective cohort study to investigate the prevalence of *M. bovis* infection in farmers (zoonotic TB) in southwest England and identify any risk factors associated with a positive result in this population. Participation in the study consisted of completing a questionnaire (supplementary information) and providing a blood sample to test for latent TB infection.

The questionnaire consisted of 32 questions, divided into four sections: participant characteristics, participant health, occupation, TB exposure, contact with cattle. The questionnaire was implemented online using REDCAP and took around 10 minutes to complete.

To test for TB infection, we used the QuantiFERON-TB Gold Plus assay (Qiagen)(9), which is the standard interferon gamma release assay (IGRA) routinely deployed in UK clinical practice for determining tuberculous reactivity. The assay is an enzyme linked immunoassay (ELISA) that measures IFN-gamma secretion by T-cells following stimulation with mycobacterial antigens (ESAT-6/CFP-10 peptides). Since these antigens are common to both *M. tuberculosis* and *M. bovis*, IGRAs using these antigens can be used to detect people exposed to either bacterium. Specificity estimates are based on low-risk populations, and are 98.9% (97.9 – 99.5)% for high thresholds(9). A blood sample was collected as per Quantiferon protocol, and an additional serum sample was collected and deposited to the Bristol Biobank for future immunological analyses.

Participants were invited to take part if they met the eligibility criteria:

1. were ≥18 years of age
2. had active involvement with TB-infected cattle in southwest England
3. no history of TB and no close contacts with TB
4. not experiencing any TB symptoms.

In this first wave of recruitment we required that participants had had at least 10 reactors in last 2 years in their herd; this was relaxed to be able to include a broader range of occupations such as veterinarians and abattoir workers.

The study was granted ethical approval by the NHS Health Research Authority Research Ethics Committee (REC) on 4 October 2021 and the amendment to expand the eligibility criteria was approved on 22 July 2022 (REC reference 21/YH/0241).

#### Target sample size

In southwest England, 48 cases of *M. bovis* were diagnosed in the five years 2015 to 2019(11), taken together with the statistic that 60% of TB cases can be cultured, and therefore genotyped(12), suggests that an additional 32 *M. bovis* cases may be treated but not identified as. *M. bovis* (80 cases in total). If 5% of LTBI cases develop active TB within 5 years of exposure, that suggests that ∼400 persons could have LTBI infection due to *M. bovis* in southwest England. To calculate the size of the exposed population, we used Defra statistics of ∼4,000 cattle herds with ‘Officially Not TB Free’ status in Southwest England. Testing one person per herd, suggests a hypothesized LTBI prevalence of approximately 10%.

Given our LTBI prevalence, 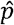, in the farmer population, the approximate 95% confidence interval will be given by 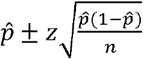, where *n* is the sample size and *z* = 1.96. If we want this confidence interval to not include a background prevalence rate of *r* = 1% (considered to be the expected background rate for LTBI in this area of England(13)), then we require

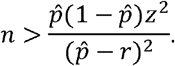

For values 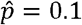, *z* = 1.96 and *r* = 0.01, the minimum required sample size is *n* = 42.7, i.e. 43 participants. Additionally, we calculated that with a low-risk group of farmers (LTBI prevalence ∼5%) and a high risk group of farmers (LTBI prevalence ∼20%) we would need 76 samples per group, i.e. 152 in total.

### 3. Study implementation

#### Study test days

Due to the practicalities of obtaining a blood sample using a trained phlebotomist and transporting the samples back to a single testing laboratory, it was necessary to conduct sampling at defined locations. We conducted three waves of recruitment at agricultural shows.

- Wave 1: 6 October 2021, The Dairy Show, Bath and West Showground, Shepton Mallet, Somerset, BA4 6QN, UK.
- Wave 2: 5 October 2022, The Dairy Show, Bath and West Showground, Shepton Mallet, Somerset, BA4 6QN, UK.
- Wave 3: 2 November 2022, Agrifest Southwest, Devon County Showground. Westpoint. Clyst St Mary. Exeter. EX5 1DJ, UK.

We conducted a fourth and final wave of recruitment on 13 March 2023 at a public house near *Bristol*. We promoted the study in advance of test days via traditional and social media, farming forums, newsletters, local agricultural shops, veterinary practices and word of mouth. On the study days, we recruited participants who were attending the agricultural shows.

### 4. Data linkage and analysis

REDCap was used to capture survey responses. Raw data was exported and cleaned in R (R version 4.0.3 (2020-10-10)).

For participants who were farmers, we collected County-Parish-Holding (CPH) number as part of the questionnaire together with consent to access the associated TB test records from the Animal and Plant Health Agency (APHA). Following each wave, CPH numbers or farm addresses for consenting participants were supplied to APHA for data extraction.

For each CPH, we calculated for the time period from 01/01/2011 to the study date: number of animals tested, number of reactors (by any test type), number of animals with lesions or culture positive at post-mortem. We characterised farms as either low, medium or high risk based on number of reactors since 01/01/2011. We defined low risk as farms/CPHs with fewer reactors than the median number of reactors, medium risk as CPHs with higher than the median but less than the upper quartile number of reactors and high risk as CPHs with a greater number of reactors than the upper quartile. Where more than one CPH was associated with a single participant, we summed the numbers of cattle tested, reactors and lesioned animals over all premises. Where more than one participant was associated with a single CPH, we used the same values for each person.

From the IGRA test results we calculated the percentage positive and the associated 95% confidence interval assuming a binomial distribution. We explored the univariate relationship between the quantitative IGRA results (TB2 – TB1 minus the Nil) and number of reactors per herd and number of reactors with lesions per herd using a generalized linear model in R with a Gaussian distribution function.

## Results

Informed e-consent was collected from 103 eligible participants; 13 participants were excluded because they completed the questionnaire online but did not attend a test date for IGRA testing. Public contributors were critical in shaping recruitment and study design. Recruitment at the dairy agricultural shows was more effective than the beef-focused agricultural show or the separate event. We recruited 26 participants in wave 1 at the Dairy Show in 2021, 41 participants in wave 2 at the Dairy Show in 2022, 18 participants in wave 3 at Agrifest, and 5 participants in wave 4 at the separate event.

Median age (IQR) (range) was 45.5 years (32-57.7) (19-77) and that did not vary between recruitment waves. A total of 52/90 participants (58%) were male and 88/90 (98%) were of white ethnicity. Due to our sampling strategy and approach, the majority of participants were farmers (68/90) or farm workers (11/90) and 11/90 were veterinary workers. Over half of participants reported working in their role for more than 20 years (53.3%).

The median household size was 4 (range: 1-6). BCG vaccination was self-reported by 59 participants (65.5%), with a median time of 14 years (range: 8-22) since vaccination to study enrolment; self-reported health status was perceived to be very good with a median score of 90/100 (range: 30-100) and most participants had not travelled to a human TB high risk area in the last 12 months (94%).

Linking participants to their APHA data allowed a comparison of reported TB burden from the questionnaire and actual TB burden. There was a positive relationship between the number of reactors reported in the questionnaire and the number recorded in the TB testing records (*R*^2^ = 0.23, fig 2). Most farmers estimated a lower number of reactors than the APHA records. Of participants with at least one reactor in the previous two years, 19 out of 66 participants (29%) reported a number that was within a factor of two of the APHA records. There was less certainty about how many of their reactors had been found with lesions at slaughter (*R*^2^ = 0.02). Eleven out of 66 participants (17%) reported a value that was within a factor of two of the APHA recorded value for lesioned animals.

**Figure 2:**
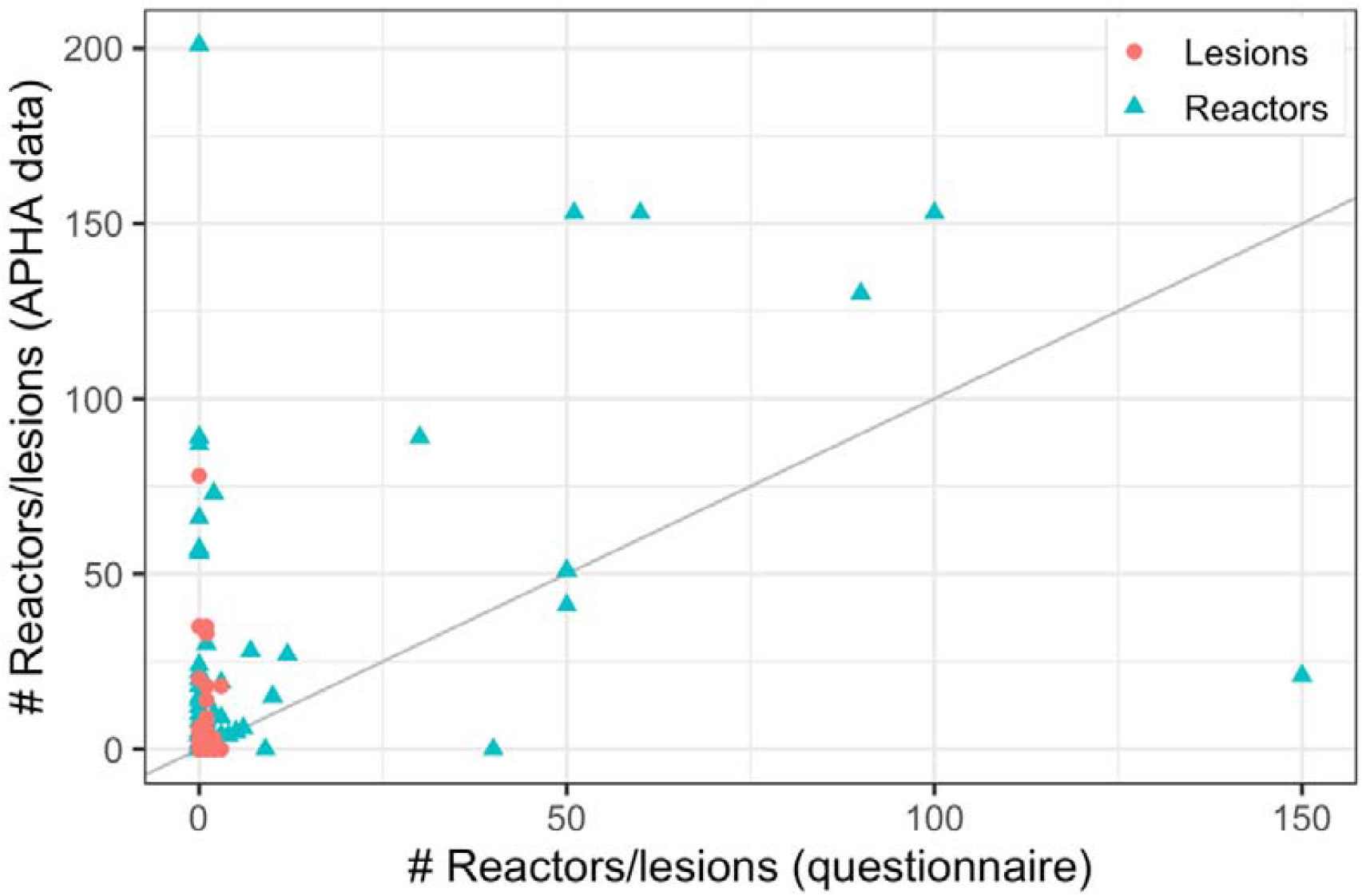
Questionnaire data versus Animal and Plant Health Agency (APHA) records. The number of reactors and number of cattle found with lesions at slaughter per farm in the previous two years as reported in the study questionnaire (horizontal axis) and in APHA test records (vertical axis).

For the remaining analysis, we used the APHA data to characterize TB burden and exposure. 76 participants were farmers or farm managers and primarily associated with a single County-Parish-Holding (CPH) number. Of these farms, the number of reactors per farm since 1 January 2011 ranged between 0 and 747. The median number of reactors per CPH was 31 reactors, and the upper quartile was 104 reactors since 01/01/2011. The low-risk farmers (<31 reactors since 2011) contained 33 participants; the medium-risk farmers (31-104 reactors since 2011) contained 23 participants and 25 participants were classed as high-risk farmers (>104 reactors since 2011). There was no difference between demographic factors (age, gender, ethnicity, BCG status) between risk groups of farmers. The non-farmers were on average younger than the farmer groups.

Herd size and number of cattle tested increased across the risk groups. The average farm had 12,800 tests conducted since 2011, ranging between 138 and 91,000. The number of lesioned animals increased with number of cattle tested and number of reactors, although there was overlap between the groups. The low-risk group had a median of two reactors with lesions (maximum 14 reactors with lesions). In the medium risk group, the median was 25 lesioned animals since 2011 and in the high-risk group the median was 124 lesioned animals since 2011, ranging from 14 to 225. 95% (77/81) of these participants reported daily direct contact with cattle.

Despite high levels of TB in their herds, not all participants considered themselves to be at risk of TB themselves, including those who we defined as high risk. There were high levels of raw milk consumption particularly in the medium and high-risk groups. Approximately half of participants in the low-risk group reported frequently consuming raw milk and 87% of those in the medium and high-risk groups reported frequently consuming raw milk. All participants in the high-risk group reported using biosecurity measures, most commonly cleaning and handwashing.

Amongst non-farmers, there were lower rates of raw milk consumption (1 in 5 participants), and they reported less frequent direct contact with cattle. Nevertheless, all of this group considered themselves to be at risk of TB infection. This group also reported high levels of precautionary measures used to prevent the spread of zoonoses.

One participant in the high-risk farmer group tested IGRA positive, all other participants were IGRA negative. The test positive participant had a TB1 minus Nil reading of 3.75 IU/ml and a TB2 minus Nil reading of 4.36 IU/ml. The remainder of the participants had TB1 and TB2 minus Nil values <0.15 IU/ml, which is below even more stringent criteria for positivity(fig 3)(14).

**Figure 3:**
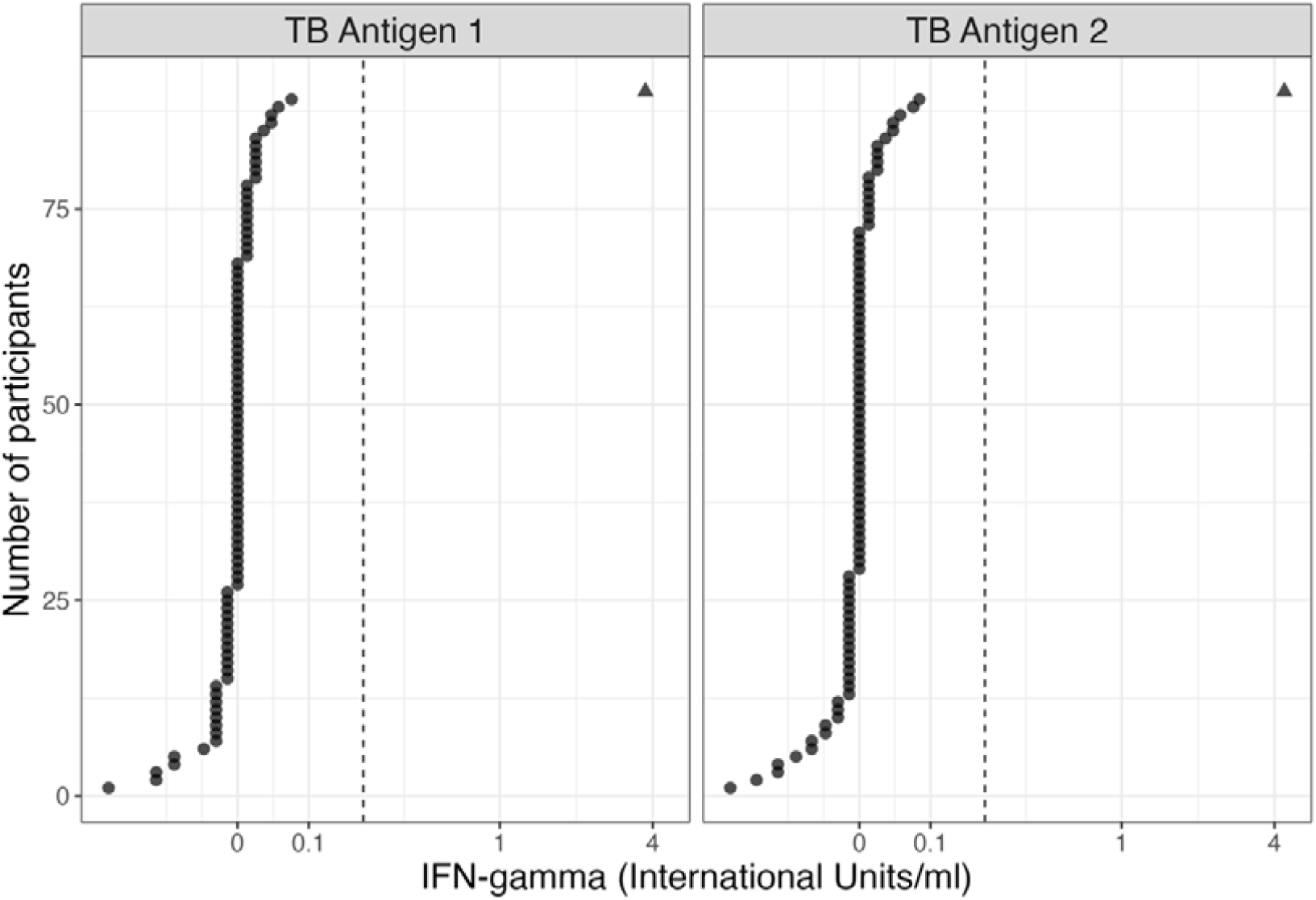
Quantitative IGRA test results for the 90 participants. The two panels are TB1 minus Nil and TB2 minus Nil. The dashed vertical line indicates the cut-off threshold for positivity(9). The positive case is indicated by a triangle, negative cases are circles.

Including all participants, the estimated IGRA positivity is between 0.05% and 7%, however this varied by risk group. All groups had an estimated IGRA positivity of 0%, apart from high-exposure individuals, in whom estimated IGRA positivity rates was 4.0% (95%CI 0.21%-22%). Considering IGRA negative participants only, we found no association between IFN-g levels and the measures of TB exposure considered.

## Discussion

In this study, we investigated TB exposure and latent TB infection (LTBI) in persons with occupational exposure to TB-infected cattle in southwest England to understand the increase in human *M. bovis* cases in England over the past 20 years. We achieved high levels of engagement from the farming community. We found high levels of potential *M. bovis* exposure, including via direct contact and consumption of unpasteurized milk. However, there was minimal evidence of LTBI infection. One out of 90 participants had a positive IGRA result, and they were classified in our group with the highest levels of potential TB exposure. Future work should focus on this high-risk group.

Building trust and engagement with the farming community was key to the success of the study. Due to the COVID pandemic, it was necessary to re-think farmer recruitment. One PPIE group suggested agricultural shows for recruitment, and this proved to be successful – on our second study day we recruited and tested 41 farmers in a single day. We worked with a regular PPIE farming contributor to develop materials and he advertised the study on farming forums and in-person at the agricultural shows. We were not able to assess the representativeness of farmers attending agricultural shows, and our sample might be biased and not reflect general biosecurity and herd health standards. Personal recruitment (either face-to-face or over the phone) was essential, and this proved the limiting factor for recruitment numbers.

Linkage across multiple data sources added strength to our results. The majority of farmer participants knew their unique County-Parish-Holding number and gave consent for linkage with their herd TB testing records. Using these linked data, we were able to get a detailed characterization of TB exposure in terms of number and type of reactors, and presence of lesions at slaughter over different time periods. We also asked about TB in their herds and found that most farmers underestimated their TB exposure. This demonstrates the viability and benefit of linking questionnaire data to testing records. Testing records could be used to target future studies or identify persons at increased risk of zoonotic infection.

The results of the study suggest that the target population was too broad, and either a higher minimum numbers of reactors was required or a larger sample size to capture more individuals with high levels of exposure. Our study design was based on knowledge of *M. tb* natural history in humans and the assumption that exposure, infection and disease with *M. bovis* is similar. The commonly used paradigm for *M. tb* is that 50% of high intensity exposures lead to infection(15). Via the questionnaire, we measured apparently high levels of potential *M. bovis* exposure in the UK, although this did not translate to IGRA positivity. Even in high burden bovine TB countries, the prevalence of *M. bovis* in bulk milk tanks was 5% (95%CI: 0%–21%) (16), therefore it might be that the infection pressure to farmers is too low to result in a positive IGRA. We hypothesize that a) TB-infected cattle are identified by the test-and-slaughter system before it is advanced enough to be infectious to humans and/or b) exposure to *M. bovis* via ingestion leads to infection with a lower probability than for *M. tb*.

It has been suggested that latent *M. bovis* infection progresses to *M. bovis* disease with a lower probability than for *M. tb* (17), although evidence is limited. The numbers of *M. bovis* cases in the UK and the suggested low rate of LTBI in individuals exposed to *M. bovis* could be consistent with a higher progression rate from latent *M. bovis* infection to *M. bovis* disease. If this were the case, it could explain the lower than expected number of IGRA positives in our study and suggests that we used an inflated effective size in our sample size calculations. Future work includes a more detailed characterization of latent *M. bovis* infection.

Other studies have measured latent TB infection in persons in close contact with cattle(18–22), almost exclusively in high TB burden settings where background LTBI prevalence is high. A study in Columbia found an LTBI prevalence of 36% in farm workers(20); a study in Mexico found a prevalence of 76.3% in dairy farm and abattoir workers and their household contacts(23); a study in Texas diagnosed LTBI in 14 out of 140 dairy workers (10%)(21). Less is known about LTBI prevalence in lower burden settings. Since our study was conducted, a UKHSA rapid review of latent TB in occupational groups in contact with cattle identified three studies in the UK from before 2004(24). Not surprisingly, given universal BCG vaccination in the UK until 2005, high levels of Heaf test positivity were found in UK farm workers. No active disease was found. Interpreting the single LTBI positive in our study, the IFN-g levels suggest that the LTBI positive case is unlikely to be a false positive(9).

In conclusion, our study found low levels of LTBI positivity in persons working with TB-infected cattle in the UK. Our results highlight the limited knowledge of *M. bovis* natural history in people and future work should include detailed follow up on *M. bovis* positive cases and contacts in order to establish progression rates from latency to active disease. Our findings could be consistent with increased LTBI risk in individuals with the highest exposure levels. Future work would need to focus on these individuals with an increased sample size.

## Data Availability

Anonymised questionnaire data for participants who consented for further research use are available to researchers upon request at https://data.bris.ac.uk/.

## Funding

The study and AT was funded by a Wellcome Trust Seed Award 217509/Z/19/Z. We received funding from the NIHR HPRU in Behavioural Science and Evaluation at the University of Bristol for PPIE. CM time is supported by the National Institute for Health and Care Research Applied Research Collaboration West (NIHR ARC West) and the National Institute for Health and Care Research Health Protection Research Unit in Behavioural Science and Evaluation.

## Acknowledgements

We would like to thank the participants for helping to shape the study and taking part.

---

**Table 1:** Demographic, TB exposure and behaviours for participants by risk group.

|  | Risk group |  |  |  |
| --- | --- | --- | --- | --- |
|  | **Low**<br>**(N=33)** | **Medium**<br>**(N=23)** | **High**<br>**(N=25)** | **No CPH**<br>**(N=14)** |
| **Age in years** Median [Min, Max] | 53.0 [20.0, 71.0] | 49.0 [19.0, 71.0] | 47.0 [27.0, 77.0] | 33.5 [24.0, 68.0] |
| **Occupation** |  |  |  |  |
| Farm Manager/Director | 1 (3.0%) | 2 (8.7%) | 3 (12.0%) | 2 (14.3%) |
| Farmer/Farm worker | 31 (93.9%) | 19 (82.6%) | 21 (84.0%) | 1 (7.1%) |
| Veterinary worker | 0 (0%) | 0 (0%) | 0 (0%) | 8 (57.1%) |
| Other | 1 (3.0%) | 2 (8.7%) | 1 (4.0%) | 3 (21.4%) |
| **Gender (Male / non-missing) (%)** | 24/33 | 12/22 | 15/25 | 8/14 |
| **Ethnicity (White British / total) (%)** | 33/33<br>(100%) | 23/23<br>(100%) | 25/25<br>(100%) | 13/14<br>(93%) |
| **BCG vaccinated (%)** |  |  |  |  |
| Yes | 23 | 11 | 19 | 11 |
| No | 8 | 10 | 4 | 2 |
| Don't know | 2 | 2 | 2 | 1 |
| **General health 1-100 scale** Median [Min, Max] | 90 [70, 100] | 90 [35, 100] | 90 [30,100] | 90 [40, 100] |
| **Number of cattle tested since 01/01/2011** Mean [Min - Max] | 4630 [138, 22800] | 12300 [2760, 77400] | 23500 [4340, 91000] | NA |
|  | Low<br>(N=33) | Medium<br>(N=23) | High<br>(N=25) | No CPH<br>(N=14) |
| **Number of reactors since 01/01/2011** Median [Min - Max] | 9.00 [0, 24.0] | 64.0 [31.0, 100] | 285 [106, 747] | NA |
| **Number of reactors with lesions since 01/01/2011** Median [Min -Max] | 2.00 [0, 14.0] | 25.0 [4.00, 50.0] | 124 [14.0, 225] | NA |
| **Consumes raw milk (frequently/non-missing)** | 16/33<br>(48%) | 21/23<br>(91%) | 20/24<br>(83%) | 3/14<br>(21%) |
| **Consider themselves to be at TB risk\* (Yes / non-missing)** | 18/26<br>(69%) | 11/14<br>(79%) | 11/13<br>(85%) | 13/13<br>(100%) |
| **Biosecurity measures to limit zoonoses spread\* (Yes/non-missing) (%)** | 18/26<br>(69%) | 12/14<br>(86%) | 13/13<br>(100%) | 12/13<br>(92%) |
| **Contact with cattle (Daily/total) (%)** | 30/33<br>(91%) | 23/23<br>(100%) | 24/25<br>(96%) | 10/14<br>(71%) |
| **IGRA result** (number of positives) | 0 | 0 | 1 | 0 |
| Negatives only: TB1 minus Nil Mean [Min, Max] | 0.00545 [-0.08, 0.05] | -0.005 [-0.05, 0.02] | 0.0025 [-0.02, 0.04] | 0.00357 [-0.05, 0.07] |
| Negatives only: TB2 minus Nil Mean [Min, Max] | -0.00576 [-0.08, 0.08] | -0.00864 [-0.06, 0.05] | -0.000417 [-0.02, 0.03] | 0.00786 [-0.04, 0.04] |
| Estimated IGRA positivity (%) [95%CI] | 0% [0%, 0%] | 0% [0%, 0%] | 4.0% [0.21%, 22%] | 0% [0%, 0%] |

*\*Question was added from wave 2 onwards.*

## Notes

**Funding statement** The study and AT were funded by a Wellcome Trust Seed Award 217509/Z/19/Z. The NIHR HPRU in Behavioural Science and Evaluation at the University of Bristol supported PPIE. CM time was supported by the National Institute for Health and Care Research Applied Research Collaboration West (NIHR ARC West) and the National Institute for Health and Care Research Health Protection Research Unit in Behavioural Science and Evaluation.

**Conflict of interest disclosure** There are no conflicts of interest to disclose.

### Competing Interest Statement

The authors have declared no competing interest.

### Clinical Protocols

https://zootb.blogs.bristol.ac.uk/study-protocol/

### Funding Statement

The study and AT were funded by a Wellcome Trust Seed Award 217509/Z/19/Z. The NIHR HPRU in Behavioural Science and Evaluation at the University of Bristol supported PPIE. CM time was supported by the National Institute for Health and Care Research Applied Research Collaboration West (NIHR ARC West) and the National Institute for Health and Care Research Health Protection Research Unit in Behavioural Science and Evaluation.

